# Implementation outputs and outcomes of a community-based maternal and newborn care model in rural Galmudug, Somalia: an implementation research study

**DOI:** 10.64898/2026.07.01.26357076

**Authors:** Naoko Kozuki, Grace Kimemia, Hassan Aden Abdi, Ahmed Abdi, Abdiwahab Maalim, Mamothena Carol Mothupi, Maryan Miris, Asia Mohamed Mohamud, Geeta Nanda, Mohamed Ahmed Omar, Mikaela Cochran-George, Derrick Machora, Muna Jama

## Abstract

Maternal and newborn health (MNH) outcomes in Somalia remain among the worst globally, driven in large part by limited access to facility-based care, particularly in rural and underserved communities; approximately one in five births take place in a health facility. To address major MNH service delivery gap in rural Somalia, the International Rescue Committee implemented a community-based maternal and newborn care (CBMNC) program, delivering a package of evidence-based interventions across selected villages. An implementation research study was conducted to generate transferable learning to inform the scale-up of comparable community-based MNH programs in similar low-resource and humanitarian settings.

The study triangulated and synthesized data from existing primary research data (e.g. population-based surveys before and after program implementation, qualitative program acceptability study, cost-efficiency analyses), program monitoring data, and program documentation. The data were organized using Proctor et al.’s Implementation Research Outcomes Framework, exploring the outcomes of acceptability, appropriateness, feasibility, fidelity, adoption, implementation cost, penetration, and sustainability. The synthesis consisted of meta-summaries across sub-domains under each outcome area.

The program delivered through 34 community health workers (CHW) served 1,165 women across a 24-month period. The program demonstrated strong acceptability among participants, who reported trust in CHWs, respect for cultural and religious norms, and tangible improvements in their pregnancy health knowledge. CHWs similarly expressed intrinsic motivation and a sense of fulfillment in their roles, despite notable challenges around workload, geographic barriers, and financial burdens. The model showed reasonable epidemiological and sociocultural fit, though some recommended health behaviors conflicted with entrenched traditional practices, such as avoiding colostrum or giving sugar water to newborns. Equity gaps were identified, particularly the underrepresentation of women with disabilities. Feasibility was constrained by household dispersal, seasonal mobility, complex task management, and an initially irregular visit schedule, which CHWs ultimately simplified to a monthly system. CHW competency improved markedly over time, with average assessment scores reaching 94% by the program’s end. Coverage was broad, with 88% of women who delivered during the program period enrolled. Sustainability considerations remained underdeveloped, representing a key area for future programming.

The CBMNC model demonstrated potential to expand access to MNH services in rural settings, with CHWs earning broad trust and acceptance among women and communities, and the intervention widely regarded as a good epidemiological, sociocultural, and contextual fit. Nevertheless, the findings make clear that achieving successful and sustainable scale-up will require more than replicating the care package itself. Practical challenges, including CHW workload, referral pathways, health literacy, and equitable reach to marginalized groups, must be deliberately addressed. Ultimately, expansion efforts must invest as much in strengthening the underlying systems that enable effective delivery as in the content of care provided.

## Introduction

Maternal and newborn health (MNH) outcomes in Somalia remain among the worst globally, driven in large part by limited access to facility-based care, particularly in rural and underserved communities (1). Approximately one in five births take place in a health facility, only one in three is attended by a skilled provider, and fewer than 10% of women receive four or more antenatal care visits (2). Women in rural, nomadic, poor households and internally displaced communities are less likely to access facility-based care than urban, educated and wealthier women (3). These access gaps contribute to a maternal mortality ratio of 621 deaths per 100,000 live births, a neonatal mortality rate of 36 per 1,000 live births, and a stillbirth rate of 28 per 1,000 births (1, 2). These levels place Somalia far from the Sustainable Development Goal targets of reducing maternal mortality below 70 per 100,000 live births and neonatal mortality below 12 per 1,000 live births by 2030 (4).

Somalia’s health policies support community-based maternal and newborn service delivery in recognition of these large gaps in service access. The Somalia Community Health Strategy 2025-2029 (5), the national Reproductive, Maternal, Newborn, Child and Adolescent Health strategy, and the national Every Newborn Action Plan (6) all identify community-based service delivery as essential for closing access gaps in rural and underserved areas. The *Marwo Caafimaad* program is the main national initiative for delivering on this policy intent. It deploys female community-based health workers, now formalized as the unified Female Health Worker cadre under the 2025-2029 strategy, to provide primary care, including MNH services, at the household level (7). In practice, however, the program has not yet been scaled to many of the rural locations it was originally intended to reach. In short, despite the enabling policy environment, community-based service delivery of maternal and newborn services is generally inadequate, and needs strengthening to address access gaps especially for vulnerable populations (8).

To address the service delivery gap in rural Somalia, the International Rescue Committee (IRC) implemented a community-based maternal and newborn care (CBMNC) program from December 2023 - December 2025, delivering a package of evidence-based interventions across selected villages. The package of interventions included MNH commodities and counseling, and was delivered through trained female community health workers (CHW). The CBMNC program was intended both as a service delivery response to the rural access gap and as a learning opportunity, designed to generate evidence on how community-based MNH services can be delivered, sustained, and scaled, including findings that would inform the operations of the aforementioned *Marwo Caafimaad* program.

Implementation evidence to guide service delivery in fragile, conflict-affected, and humanitarian settings remains scarce, and recent calls have highlighted the need to expand implementation science in such contexts (9, 10). Implementation research generates practice-oriented evidence on how programs work in real-world settings, including what works, what does not, and what adaptations are needed for sustained delivery and scale-up (11, 12). The objectives of this study were to synthesize evidence on the implementation of the CBMNC program across the eight Implementation Research Outcomes Framework domains of acceptability, appropriateness, feasibility, fidelity, adoption, implementation cost, penetration, and sustainability and generate transferable learning to inform the scale-up of comparable community-based MNH programs in similar low-resource and humanitarian settings.

## Methods

This study is a triangulation and synthesis of multiple data sources generated during the implementation of the CBMNC program in rural Somalia, organized using the Implementation Research Outcomes Framework (13). The synthesis draws on primary research, routine program monitoring and evaluation outputs, and program documentation, to be described below.

### Program description

The CBMNC program was implemented by the IRC in partnership with the Federal Ministry of Health and the Galmudug State Ministry of Health in Dhusamareb district, Galgaduud region, Somalia. Galgaduud is in central Somalia within Galmudug State. Dhusamareb functions as the administrative capital for both Galmudug State and the Galgaduud region (2). The district includes Dhusamareb town and surrounding rural settlements, where households predominantly depend on pastoralist and agro-pastoralist livelihoods and live in dispersed communities. Rural populations in this region face persistent barriers to accessing MNH care, including long distances to health facilities, limited transportation, seasonal migration, displacement due to drought, and insufficient connections between health facilities and communities (14).

The CBMNC program was established as a pilot intervention to address MNH in rural areas with limited access to facility-based care. This implementation research study was originally conceptualized in 2020, aimed to examine the anticipated expansion of the *Marwo Caafimaad* program within a few years of the study conceptualization. However, *Marwo Caafimaad* had not reached the targeted rural locations at the study’s outset and remained absent at the time of writing in mid-2026. Subsequently, the IRC received funding to implement a community-based service delivery model for MNH, which was then designed to closely align with the scope and objectives of *Marwo Caafimaad*. This alignment was intended to ensure that lessons learned from the pilot and the implementation research study could inform the future scale-up of nationally-aligned community-based MNH service delivery models.

The program’s primary objective was to increase uptake of essential newborn care practices (early initiation of breastfeeding, thermal care, clean cord care) through household-level counseling, and as secondary objectives, increase uptake of other behaviors, such as uptake of key commodities and identification and seeking care for danger signs. Service delivery occurred through home visits during pregnancy and the postnatal period. More details are available in Table 1. The intervention package was determined based on stakeholder consultations and constrained optimization analyses, considering aspects of each intervention such as expected mortality impact, CHW deliverability, commodity availability, cost, and national policy permissions. Details of the process are available elsewhere (15).

**Table 1:**
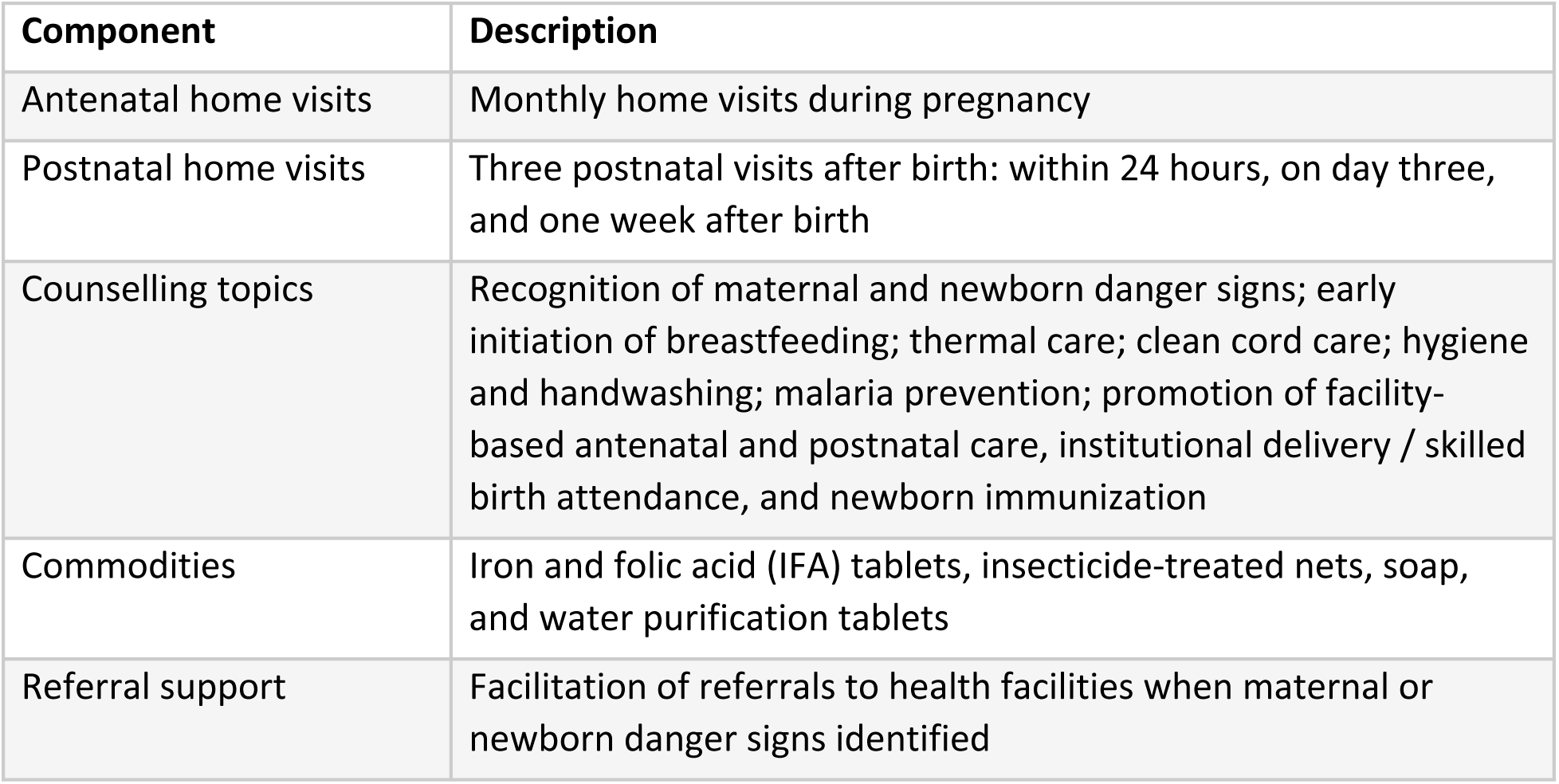
Community-based maternal and newborn care service package delivered by CHWs.

The CBMNC program operated from December 2023 to December 2025 in eight remote villages within Dhusamareb district: Gadoon, Balicad, Mareerguur, Ceeldheere, Faragooy, Halanley, Haadfull, and Olol. These villages are situated approximately 20 to 65 kilometers from the nearest functional health facility, presenting significant barriers to both routine and emergency maternal and newborn health care. The program specifically targeted pregnant women residing in these catchment areas. A one-time pregnancy mapping exercise identified pregnant women in the relevant catchment areas at the start of the program, followed by rolling identification and enrollment by CHWs through household visits and word-of-mouth.

At any point in time, a total of 34 trained female CHWs from the target villages delivered services, supported by supervisors and Ministry of Health liaison officers. The CHW cadre included women from the local communities, ranging from young adults to older women with established community roles. Most CHWs were married, all were literate in Somali, and the majority had at least a primary education. Training covered the Federal Ministry of Health community health curriculum, the CBMNC intervention package, data and reporting tools, commodity management, safeguarding, communication, and referral processes. Recruiting female CHWs from the same villages was intended to enhance trust, acceptability, continuity of contact, and access to women in remote rural settings. National and state stakeholders helped shape and guide the program. A federal-level Technical Advisory Group, co-led by IRC and the Federal Ministry of Health, brought together government, United Nations agencies, donors, and implementing partners to review progress, address challenges, and support alignment with national and state MNH priorities. The home-visit model was complemented by structured community engagement activities, summarized in Table 2.

**Table 2:** Community engagement strategies used in the CBMNC program.

| Activities | Purpose and delivery |
| --- | --- |
| **Mother-to-mother and father-to-father support groups** | These groups reinforced MNH messages, facilitated peer learning, and promoted household support for recommended practices. Mother-to-mother groups were organized separately for mothers under 18 and for older mothers to ensure safer, age-appropriate environments for open discussion of pregnancy, childbirth, and newborn care. |
| **Engagement during existing community health committee meetings and other community stakeholder dialogues** | The program engaged existing community structures to support community ownership, facilitate problem-solving, promote acceptance of CHWs, encourage referrals, and enable discussion of local concerns. Participants comprised of village leaders, religious leaders, traditional birth attendants, women’s representatives, and other influential community members. |
| **Community-based health education sessions** | Sessions with the general public to expand awareness of antenatal and postnatal care, maternal and newborn danger signs, essential newborn care, and facility care-seeking. Visually adapted, Somali-language materials were made available. |

### Research framework

We identified potential implementation research frameworks through review of widely cited implementation science theories, models, and frameworks, and assessed them against published criteria for framework selection that consider a project’s purpose, scope, available data, and resources (16). The Implementation Research Outcomes Framework (13) was selected because its evaluative orientation matched the purpose of the synthesis, which was to characterize the implementation of the CBMNC program across multiple implementation outcomes. This framework defines eight outcomes: acceptability, adoption, appropriateness, feasibility, fidelity, implementation cost, penetration, and sustainability. (See Table 3 for definitions.) These implementation outcomes are distinct from service outcomes, which will be reported in the future in a parallel analysis of pre- and post-program implementation surveys, although some reflections from those data will be incorporated here. The study team adapted the framework by creating subdomains to allow for more nuance in interpretation of each outcome, with some sub-domains expanding the original definition by Proctor et al. Sub-domains under the acceptability outcome were drawn from the Theoretical Framework of Acceptability (17). For the remaining outcomes, sub-domains were developed by the research and program implementation teams to capture dimensions of program model relevant to the rural Somalia context. Specifically for the outcome of sustainability, it was organized as a sub-domain in each of the other outcomes to see the interaction between the select outcome and sustainability of components and findings within those outcomes.

**Table 3.** Implementation Research Outcomes Framework: outcomes and definitions.

| Implementation outcome | Definition, reproduced from Proctor et al. (13) | Subdomains* (introduced by the study team) |
| --- | --- | --- |
| Acceptability | The perception among implementation stakeholders that a given treatment, service, practice, or innovation is agreeable, palatable, or satisfactory | Affective attitude, burden, ethicality, intervention coherence, opportunity costs, perceived effectiveness, self-efficacy** |
| Adoption | The intention, initial decision, or action to try or employ an innovation or evidence-based practice. Adoption also may be referred to as 'uptake' | Supply side (motivation to deliver, CHW enrollment, retention, and role uptake, perceived role fit, barriers to role uptake, discontinuation); demand side (motivation to participate, participation patterns, initial uptake, barriers); program strategies (enablers for adoption, community engagement, policy/system support, local adaptations, feedback mechanisms) |
| Appropriateness | The perceived fit, relevance, or compatibility of the innovation or evidence-based practice for a given practice setting, provider, or consumer; and/or perceived fit of the innovation to address a particular issue or problem | Problem alignment, geographic/seasonal fit, provider capacity fit, sociocultural fit, policy/system fit, context-specific factors, equity factors |
| Feasibility | The extent to which a new treatment, or an innovation, can be successfully used or carried out within a given agency or setting | Supply side (workload, supply chain, supervision, CHW capacity, funding and cost, opportunity cost), demand side (participation, commodity uptake, behavioral uptake, physical access), contextual factors (geography and transit, insecurity and shocks, policy environment, seasonal patterns, community alignment) |
| Fidelity | The degree to which an intervention was implemented as it was prescribed in the original protocol or as it was intended by the program developers | CHW output, CHW process, supervisor contribution, barriers and adaptations |
| Implementation cost | The cost impact of an implementation effort. Implementation costs vary according to (1) the complexity of the intervention, (2) the complexity of the implementation strategy, and (3) the setting | N/A |
| Penetration | The integration of a practice within a service setting and its subsystems | Community integration, health facility integration, governance and ownership |
| Sustainability | The extent to which a newly implemented treatment is maintained or institutionalized within a service setting's ongoing, stable operations | N/A |

\*Sustainability was added as a subdomain across all outcome areas.
\*\*Taken from Sekhon M, Cartwright M, and Francis JJ (17).

### Data sources

The data triangulation and synthesis drew on sources collected from the program implementation period and a few months prior (September 2023 to December 2025), grouped into three categories: primary research conducted alongside the program, routine monitoring and evaluation (M&E) generated during implementation, and program documentation produced for internal management and reporting. Primary research components were the pre-post population-representative survey, a qualitative study on the acceptability of the program, and the cost-efficiency analysis. The first two studies were conducted in partnership between Somali Research and Development Institute and the IRC, with the former executing all data collection; details will be available in separate publications in the future. The cost-efficiency analyses methods used are available elsewhere (18). M&E outputs and program documentation were generated by the program implementation and M&E teams as part of routine program operations. Table 4 summarizes each data source by category, purpose, and outcomes it informed, and more details including data type, collection method, the team responsible for collection, period of collection, and scale or scope are available in S1 Table.

**Table 4:**
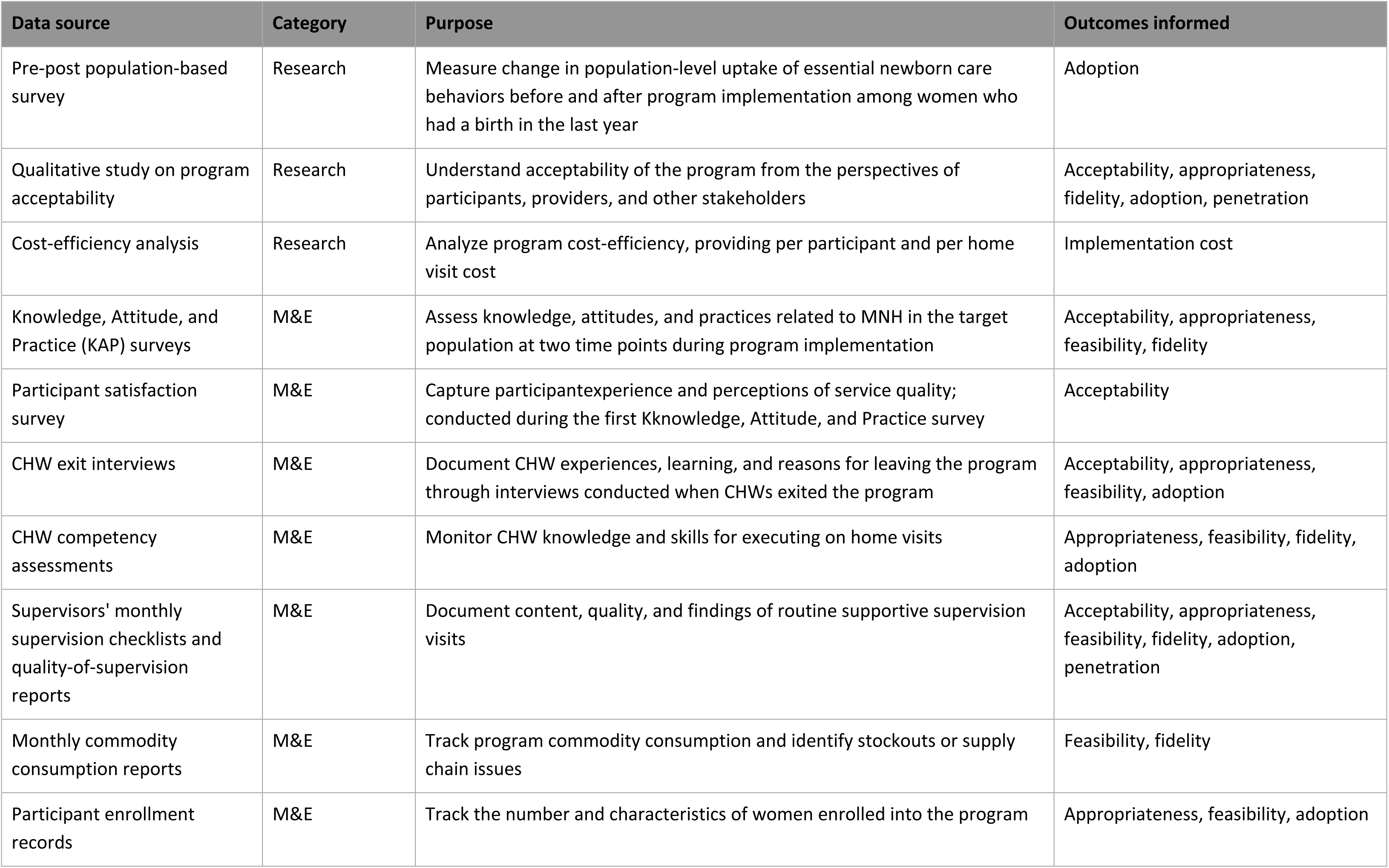
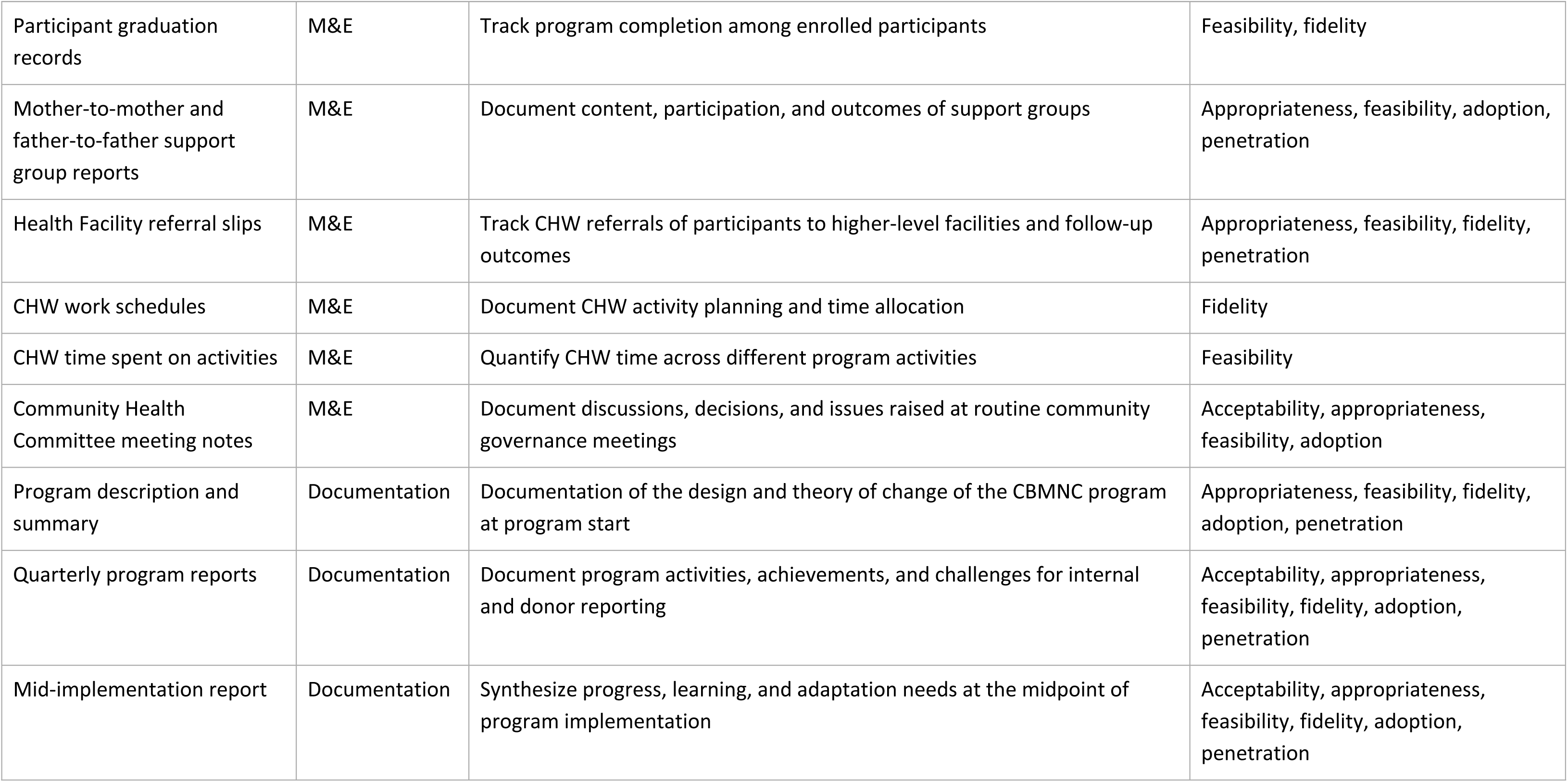
Data sources contributing to the implementation research synthesis.

### Data extraction and synthesis

We conducted data triangulation using an integrative triangulation matrix developed in Microsoft Excel, mapping data sources to key framework components. A triangulation protocol (19) was developed to guide the process, outline key objectives, procedures and definitions for the synthesis. Each outcome of the framework was allocated its own Excel worksheet and each worksheet contained columns representing outcome subdomains and rows as individual data sources.

Assigned team members extracted data from each source listed in Table 4 into the relevant subdomain/data source cells in the matrix. Both qualitative and quantitative data were extracted narratively, with excerpts used where necessary. Each subdomain was then triangulated across data sources, with the narrative summary highlighting key findings as well as a convergence analysis and program implications. Convergence analysis assessed agreement across data sources in terms of defined convergence, complementarity, divergence or silence. The team met regularly throughout the extraction period to discuss ambiguous content and support consistency across sources. Program and research team members performed the extraction (MJ, HAA, AM, MM and GK), while two separate researchers (NK and MCM) conducted the narrative meta-inference and convergence analysis. The team reviewed and discussed the completed matrix to confirm that the extractions, convergence codes, meta-inferences, and implications were internally consistent and adequately supported by the underlying source material. While reliability or member checks were not conducted on the extraction, the narrative summaries were discussed collectively and data revisited as relevant. Where thematically similar content appeared under multiple subdomains, it was consolidated under a single domain to minimize redundancy. The meta-summaries across subdomains were used as the basis of the manuscript. Additionally, the program implementation team drafted program implications based on those summaries, which partially contribute to the themes to be discussed in the discussion section of this paper.

Data quality varied across sources by purpose and method of collection. Primary research followed study protocols with trained data collectors, standardized instruments, and Institutional Review Board approval. Routine M&E outputs were generated under standard program procedures, with variable completeness across time periods and sources. Program documentation reflected internal reporting at specific time points rather than continuous data collection. During extraction, team members noted instances of incomplete records or limited coverage of a subdomain, and these were reflected in the convergence coding; sources with partial coverage of a subdomain contributed to a complementarity or silence rating rather than a convergence rating, to avoid overstating the strength of evidence.

### Research ethics

All primary research referred to here received Institutional Review Board approval from the Somalia Ministry of Health, Somali Research and Development Institute, and the IRC respectively. For the M&E data sources, the data were either extracted in an already-analyzed form by the program M&E team, or if the data were further analyzed using the original source data, no identifying information was made available to the analysis team.

### Reflexivity

The analysis team consisted of researchers and program implementers based in Somalia, Kenya, and the United States, bringing diverse perspectives shaped by professional roles, geographic positions, and direct and indirect involvement in program delivery. Recognizing that these mixed experiences could shape interpretation, we adopted procedures to separate opinion from data throughout the analysis. Prior to the analysis, all team members were trained on the study framework and the triangulation protocol, which was modified based on the team input. This ensured shared understanding of tools and processes used for extraction, triangulation, and synthesis. The tools were tested to ensure suitability for members of the team at different research skill levels.

Trained researchers provided oversight of the process, reviewing the extraction to ensure summaries were based on data sources and not anecdotes or implementer experience. However, implementer perspectives were also captured in notes and under program implication cells, to ensure a reflective process in the data analysis. Conversely, research outputs such as the narrative summaries were reviewed by implementers for sense-checking and to inform program relevant conclusions. The two-step process of researcher-led meta-inference followed by implementer-led program implications was designed so that interpretation and operational recommendations were each generated by the team members best positioned to produce them, while maintaining a clear separation between what the data showed and what the program team concluded from it. Through these procedures, the diverse experiences of the team became a strength of the synthesis without compromising the data-driven integrity of the analysis.

## Results

### Acceptability

#### Client perspective

CBMNC participants (referred henceforth as participants) reported trust in and feeling “safe” with the selected female CHWs, and not needing to travel far distances as women provided a sense of security. According to participants, the CBMNC program delivered interventions in a way that respected religious, ethical, and social norms around privacy and confidentiality; participants described confidentiality as consistent with Islamic values of trust and protecting private information. The KAP surveys conducted with participants reported high confidence in receiving MNH care from CHWs (e.g. 75% “very confident,” 21% “confident”) and that CHWs communicated MNH information adequately. Participants reported concrete positive changes; for example, participants reporting improved pregnancy health and gaining new knowledge, reinforcing their belief in program effectiveness. The initial acceptance of the program may have differed by age; young pregnant women occasionally appeared hesitant about enrolling but accepted after privacy assurances by CHWs.

Participants did not report a huge burden to participation. CHWs often provided advance notice of their home visits to ensure minimal interference with participants’ daily responsibilities. However, participants reported dissatisfaction with discontinuation of water purification tablets that occurred halfway into the program; the program team decided to discontinue the intervention altogether due to concerns about financial sustainability of a relatively expensive product. They also reported concerns about the program period only covering a short duration postpartum, which women perceived as a period of high demand for support and services. Data from one of the quarterly reports also suggested that other pregnant women’s needs such as nutritional interventions were perceived as important by community members but not covered by the program. While program scope was communicated to participants, other expectations related to comprehensive MNH care (e.g. facility-based quality of care) led to incorrectly attributed negative perceptions on the model. Routine meeting notes and reports showed gradual improvements in community attitudes over time, especially related to cultural acceptance and utilization of commodities.

#### Provider perspective

CHWs reported intrinsic value of the role, such as empowerment and personal fulfillment. However, CHWs also perceived a heavy burden in delivering the services, such as commute times, tough geographical terrain, and financial costs, including those associated with social expectations of visitors after a birth of a baby bringing gifts (*diiqista)*. Due to these expectations, some program adaptations were made; part way through, the program provided soap and cloth (*dira)* to the CHWs to gift mothers at the first postnatal visit. CHWs expressed confidence in their ability to deliver services, citing external inputs (e.g. training and tools provided) and intrinsic factors (e.g. gaining life-long knowledge). There was some data divergence around CHWs stating that they could balance the demands of the job with family responsibilities while also voicing concerns about their workload.

Facility-based health providers who were not directly involved in the model expressed positive attitudes, driven by the clear roles defined for CHWs and perceived increased uptake in facility services. However, while data from the baseline and endline surveys showed an absolute increase in institutional delivery before and after the program, no statistically significant association existed between program enrollment and facility service use after adjusting for confounding variables.

#### Supporting environment

As described in Table 2, there were several activity pathways for encouraging a supportive environment for participants. The program established 32 mother-to-mother and 14 father-to-father support groups, each comprising approximately 10 members, reaching about 460 group participants. 42 Community Health Committee engagements and two community stakeholder dialogues were held, and a total of eight quarterly community-based health education sessions were held. The data were silent on which of these pathways appeared to be the most influential in supporting community acceptance of the program. Community elders expressed initial concerns and suspicions given their unfamiliarity with the program, although periodic program reports appeared to signal that these concerns reduced over time, likely due to repeat exposure.

### Appropriateness

#### Epidemiological fit

The delivered services were selected through a statistical modeling approach called constrained optimization (process already described in program description section) (15). This exercise attempted to ensure epidemiological and policy appropriateness of the program model to context. This also meant that interventions that were deemed epidemiologically impactful due to available national and global epidemiologic evidence on causes of mortality, such as advanced distribution of misoprostol for prevention of postpartum hemorrhage or oral antibiotics for newborn sepsis, were excluded due to policy restrictions for community-based distribution. Data from the synthesis was silent on epidemiological fit as perceived by the community and other stakeholders; there were no references in other data sources around the community’s perceived causes of maternal and neonatal mortality or morbidity and if and how the model addressed those causal pathways.

#### Provider capacity

Data sources reported fit between CHW capacity and the model, though this was contingent on sustained supervision and active support from program staff to troubleshoot the piloted model. Early model adaptations were made such as simplification of data collection forms to ensure better alignment with capacity and translation of globally available MNH video training modules into the local language of Somali. Routine quarterly capacity assessments conducted on CHWs indicated consistent high performance among some, while others gradually improved over time (more in subsequent section). Supervision and targeted support further enhanced CHWs’ use of job aids, data collection tools, and participant engagement. Nine months into the program, Whatsapp groups were introduced to CHWs, their supervisors, and select program staff to have additional touch points between monthly team meetings to support capacity improvement and troubleshooting.

#### Distance and mobility

The model addressed poor access to care in remote and rural areas by selecting locally-based CHWs. Free transport support was offered to participants when referred for danger signs but did not seem to be consistent or sufficient, as some participants still reported the transport and referral cost as a barrier to accessing facility-based care according to the KAP survey. No systematic adaptations to the model were instituted to account for seasonal mobility of participants and potentially the CHWs themselves, or general challenges to CHWs operating under challenging weather patterns. There were some informal solutions proposed such as making home visits in the morning or evening to avoid heat waves. The quarterly program reports also showed limits in seasonal and geographical fit, demonstrating a drop in client enrollment (up to 25%) during the hot, drought seasons, implying both challenges around seasonal mobility and relocation of participants or CHWs themselves as well as the physical challenge of CHWs making home visits during such times.

#### Sociocultural fit

Some counseling on MNH behaviors did not align with existing cultural practices and beliefs. For instance, while breastfeeding was culturally and religiously considered important, exclusive breastfeeding held some friction with common practice, as did newborn bathing and thermal care. The data across multiple sources suggest that taking up the recommended behaviors over some of the traditional beliefs such as giving sugar to newborns or avoiding colostrum evolved over time. However, there were other behaviors that the program attempted more indirectly to promote but observed no change, such as institutional delivery, early disclosure of pregnancy among young mothers (to allow for earlier service provision), and men’s active participation in pregnancy care.

#### Equity

The model reduced some access barriers by removing financial cost and bringing services closer to rural households. Across data sources, free service provision was consistently valued, with participants and family members emphasizing that poor households could not otherwise afford commodities or health care costs. However, evidence was limited on whether the model specifically identified or reached more marginalized groups; the model did not have explicit strategies to identify and include women with disabilities or pastoral households. The capture of women with disabilities in the community, using the Washington Group questionnaire, was significantly lower in the baseline and endline surveys than the prevalence expected in the general population and from national data, limiting interpretation of program reach for this group. This could be driven by lower self-report due to stigma-related under-reporting or poor measurement in administering disability questions. There was also some silence across the data sources, limiting how far program reach and inclusion can be assessed across vulnerable groups.

### Feasibility

#### Visit schedule

CHW visit frequency and schedule were originally developed with feasibility considerations in mind; the original schedule included six antenatal visits (0–3 months, 4–5 months, 5–6 months, 7 months, 8 months, and 9 months) and three postnatal visits (within first 24 hours, 3 days, and 7 days after delivery). During implementation, two issues emerged. First, the number of mothers and visits assigned to CHWs was reported as a high burden by CHWs, particularly when taking distances and lack of transport into consideration. This was even after the model was shifted from the initial plan of one CHW to 150 households, the ratio promoted by the national *Marwo Caafimaad* program, to one to 50 after observing the dispersion of households and feasibility of visits. Second, after the first three months of implementation, the CHWs indicated that the irregularity of the antenatal visit schedule was too complicated to track. This was further complicated by other tasks such as identification and enrollment of new pregnancies and distribution of IFA; IFA was distributed first as a 30-day dose, and another 60-day dose upon completion, with the distribution of the 60-day dose not necessarily aligning with when the next visit would be. The program team with input from the CHWs eventually changed the antenatal visit schedule to monthly for scheduling ease and timely distribution of IFA, even if meant a higher workload for CHWs.

#### CHW working conditions

CHWs’ ability to reach households depended heavily on travel conditions and the dispersion of homes. CHWs tried to pre-plan their visits through phone calls to minimize inefficiencies such as making long treks when the client was unavailable; airtime that was provided by the program to CHWs was seen as critical for access. CHWs also reported a notable challenge and increased workload in trying to track women who relocated. Many CHWs themselves were pastoralists, where daily life was characterized by mobility, harsh living conditions, and competing responsibilities such as childcare, household duties, and livelihood activities, making it challenging to sustain their work. With that said, only two CHWs out of 34 dropped out of the program, both for relocation outside of the community. In villages with challenging patterns of household dispersion, the CHWs adapted by rearranging schedules to reduce total frequency of visits to specific locations within a village and on occasion arranging with the participants to find a central location for meet-up rather than making the full way to their homes.

CHWs were allotted parental leave per federal MoH policy, with about a third of CHWs taking parental leave at some point in the program period. The policy itself does not dictate arrangements for parental covers, so the program made adjustments by shifting tasks where possible or relying on a pool of reserve CHWs; the program used a pool of two to three pre-trained reserve CHWs in each village to provide short-term cover, whether for parental leave or otherwise.

#### Supply chain

The program supply chain included the following: commodities for participants (water purification tablets, IFA, Long-Lasting Insecticide-Treated Nets, soap) and supplies for the CHWs (documentation: register, monthly summary sheets, birth outcome sheet; umbrella; boots; water bottles). The supply chain followed the pathway of IRC office to supervisors to CHWs to participants. Supply chain was mostly functional throughout the implementation period, with the main notable source of disruption being the delivery of commodities from the nearest IRC office in Dhusamareb town to the CHWs in the farthest villages (approximate distance of 65 km), with one dedicated vehicle unable to keep up with demands of stock distribution on certain occasions. There were also circumstances in which the demand exceeded stock. There was one reference to a health facility staff noting that they occasionally refer their clients back to CHWs for services such as IFA, water purification tablets, and health education when the facility was out of stock or could not absorb the workload, which was not a part of the model design.

#### Supervision

The model included multiple touch points between program supervisory functions and the CHWs. For instance, quarterly supportive supervision and competency assessments for each CHW (involving a checklist of steps expected of the CHW on a home visit) were planned. Supportive supervision involved reviews of CHW data and registers, direct observation and problem-solving, agreement on action points, and recognition of CHW achievements; it also triggered refresher training or on-the-job support as well as tool and process improvements. In total, six rounds of competency assessments were held across the 24-month program period. Monthly meetings between CHWs and the program staff were also held, which improved quality and reduced errors in data and tool use. These monthly meetings helped bridge direct supervision gaps, as supervision could be inconsistent due to access. Around nine months into the program, remote support (e.g. WhatsApp group of program team, supervisors, and CHWs, phone calls or voice messages between supervisor and CHW) was launched as an additional mechanism, with monthly meetings being raised as too infrequent in touch points. The data remained silent on which of these mechanisms were most or least effective in sustaining the performance and retention of CHWs and the corresponding time burden on program staff and CHWs themselves. The program also did not address, and the data did not speak to, the quality of or CHW satisfaction with supervision.

CHW competency scores improved over time, with the capacity assessments demonstrating that by the third assessment conducted (first assessment conducted in March, second assessment conducted in July, and third assessment conducted in October 2024, relative to program start in December 2023), every CHW had met the pre-determined scoring threshold of 80%. By the last competency assessment which was conducted 23 months into the program, the average score of the CHWs was 94%. See Figure 1 for change in average score over assessments.

**Figure 1:**
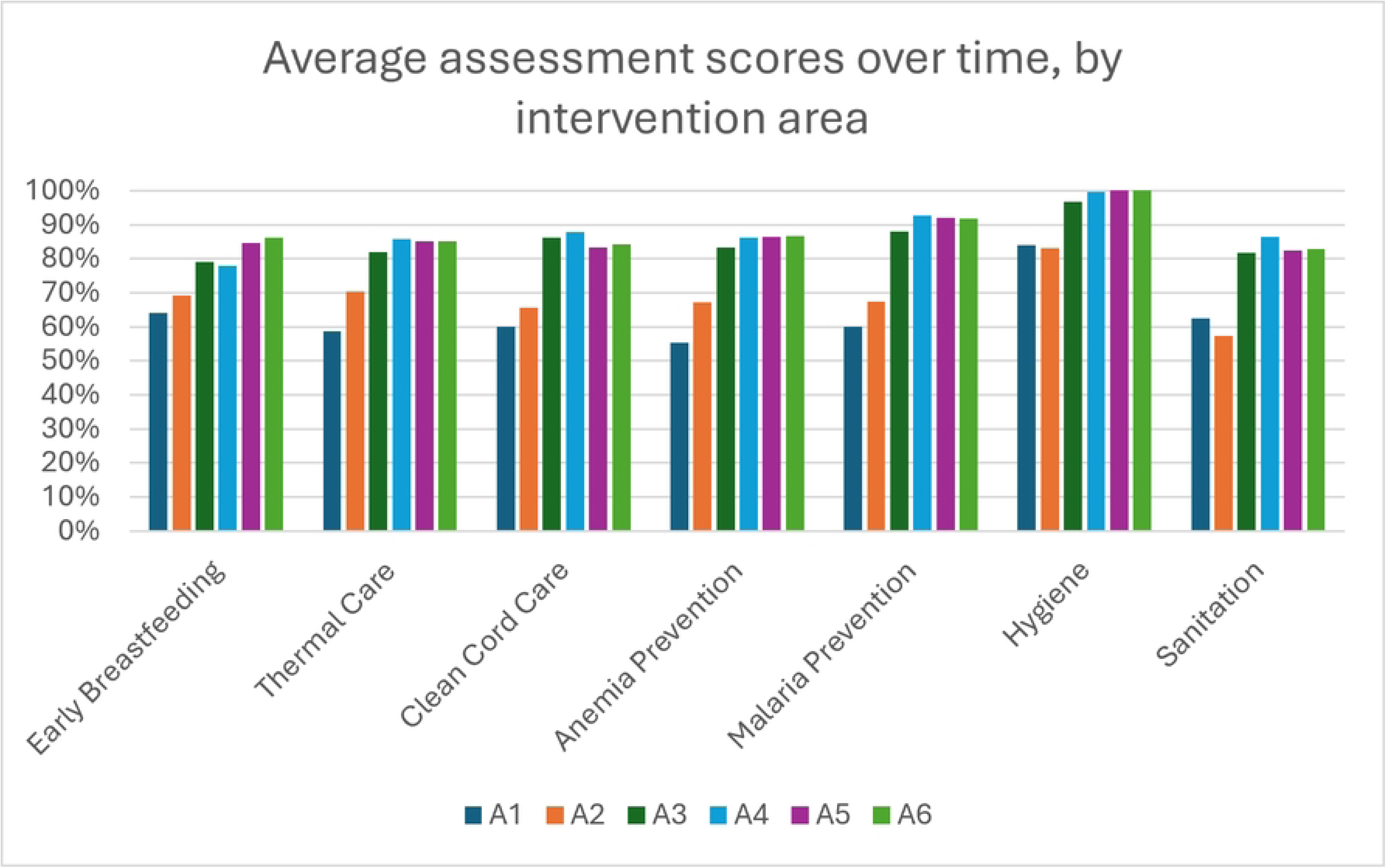
Average CHW competency assessment scores, over six quarterly assessments. *The “A” in the key (e.g. A1) is “Assessment.”

#### Contextual factors

The program monitored security situations through direct communication with local authorities. No security incidents were reported during the implementation period. There were no major changes to relevant policymakers at the state level over the duration of the program and the program maintained engagement with the state MoH and the national Technical Advisory Group. Seasonal droughts and successive failed rainy seasons intensified environmental stress, reducing both agricultural and pastoral productivity. The failed seasonal rain caused relocation where some villages were badly affected, leading to participants missing services and some of the community activities delayed. The particular overlap of high heat periods and fasting during Ramadan were also noted as a challenge in the first year, and adaptations such as provision of umbrellas for heat and advanced distribution of commodities prior to Ramadan were made for the second year.

#### Referral completion

The program significantly reduced service access issues for women residing in remote areas, with all communities served being located more than 30 km from the nearest functional health facility. However, the challenge of referral completion remained a salient theme throughout the program period, even with subsidized transport. Noting that referrals were not a major objective or an investment point for the model, arranging transport, time spent traveling and waiting at the health facility, weak facility readiness, low participant confidence in referral facilities, and relevant opportunity costs incurred throughout (e.g. missing work and other childcare responsibilities) remained too high of a barrier.

### Fidelity

#### Fidelity to process

As noted above, competency assessments demonstrated that adherence to protocol and processes for CHWs improved over time and some deviations to original protocol were executed to meet contextual demands.

#### Fidelity to intended visits and coverage

The population representative endline survey reported that 88% of women who delivered a child in the last 12 months had been enrolled in the CBMNC program. Of the total, 5% was younger than 18 years old. Given that women who were in the second or third trimester at the start of the program could not be enrolled any earlier, the data when removing the first six months of enrollment show that 53% of women were enrolled in their first trimester, 35% in their second trimester, and 11% in their third trimester, demonstrating early capture of pregnancies in the community. The same subset of women received a median 5 antenatal visits (range 1 to 8, IQR of 3-6) and a median 1 postnatal visit (range 0 to 3, IQR of 0-3). (For just the women enrolled in the first six months, the trimester breakdown was 43%, 45%, 12%, and no difference in the visit statistics.) When removing the first six month of enrollment, 16% of women received the originally intended nine total visits (109 out of 670) and 38% of women received the originally intended six antenatal visits (252 out of 670). 27% of all enrolled women did not receive a single postnatal contact.

In total 1165 women were served and 5335 antenatal and 1780 postnatal visits were made by 34 CHWs. The median of the average number of visits per month was 8. Table 5 shows the summary of the total number of clients, ANC visits, PNC visits, and total number of visits, as well as the average number of visits per month of each CHW, stratified by length of service.

**Table 5:** Number of clients enrolled and home visits made.

|  | All CHWs (n=36*, including two individuals who left the program early and two individuals who joined in their stead) | CHWs who served at least 20 months (includes those who went on parental leave) (n=32) | CHWs who served the full 24 months (excludes those who were on parental leave or relocated for other reasons) N=20 |
| --- | --- | --- | --- |
| Number of clients served | Median: 28.5<br>IQR: 17.5-48<br>Range: 8-73 | Median: 30<br>IQR: 19.5-49.5<br>Range: 14-73 | Median: 26<br>IQR: 18-46.5<br>Range: 14-73 |
| Number of ANC visits | Median: 133.5<br>IQR: 77.5-221.5<br>Range: 29-339 | Median: 137<br>IQR: 88.5-229.5<br>Range: 60-339 | Median: 121<br>IQR: 76-221.5<br>Range: 60-339 |
| Number of PNC visits | Median: 38.5<br>IQR: 26.5-73.5<br>Range: 14-117 | Median: 50<br>IQR: 30-75<br>Range: 14-117 | Median: 47.5<br>IQR: 27-70.5<br>Range: 14-117 |
| Number of total visits | Median: 171<br>IQR: 106.5-292<br>Range: 44-456 | Median: 178<br>IQR: 115.5-304<br>Range: 74-456 | Median: 158.5<br>IQR: 103-292<br>Range: 74-456 |
| Average number of visits per month | Median: 7.9<br>IQR: 4-13.3<br>Range: 3.1-19 | Median: 8.2<br>IQR: 5.5-13.5<br>Range: 3.1-19.0 | Median: 6.6<br>IQR: 4.3-12.4<br>Range: 3.1-19.0 |

\*Only 34 were operational at any single time.

### Adoption

#### CHW facilitators and barriers

Motivations indicated by CHWs included wanting to support and share knowledge with others, being inspired as part of religious practice, and personal desires for professional and personal development. Motivation was also cultivated by regular supervision touch points including recognition of achievements during monthly meetings and direct communication with the project team through a WhatsApp group. The program provided small operational enablers, such as airtime and protective shoes to support CHW motivation. Hypothesized demotivating factors include disruptions to the CHW’s tenure in the program (e.g. parental leave, joining as a replacement CHW), mismatch of expectations like community expectations that CHWs can provide more than they can be equipped to, and those already mentioned, such as workload and geographic barriers to accessing households.

#### Participant uptake

Participant participation in home visits appeared dependent on two factors: competing priorities with daily errands (e.g. household chores, market days) and individual characteristics (e.g. first-time mothers hesitating to reveal their pregnancies early). CHWs observed that first-time mothers and those in early pregnancy stages engaged with them differently than other pregnant women. In order to better support this group, the program team organized M2MSG specifically for adolescent and first time mothers, in addition to other processes that CHWs were using to ease the burden of participation, such as pre-calling before a visit, flexible rescheduling, and following up with those who missed visits that was used for all women. Commodity distribution was deemed essential by CHWs, with one noting “some mothers would not listen without these supplies.”

Generating demand for the program required significant effort, particularly in rural areas, where persistent awareness campaigns were needed to gain both mothers’ and husbands’ support. Mothers-in-law played a key facilitative role by encouraging daughters’ participation, while providers noted that uptake improved when recipients were educated about services in advance by trusted community figures such as elders and religious leaders. Some husbands and older men in the community expressed concerns around the qualification of unmarried CHWs, requiring ongoing efforts to build trust and secure family buy-in. Community engagement mechanisms were seen by program teams as promoting information sharing and broader adoption of MNH practices. However, participation data in such groups were not captured systematically and no demand-side respondents directly referenced these groups as a driver of their uptake.

Several themes emerged around specific interventions. A recurrent challenge described by some clients was adhering to IFA supplementation, mainly in relation to side effects but also in remembering to take the pills. For instance, in one KAP survey conducted earlier in the program period (eight months into the program), IFA adherence experienced a notable drop-off: 36.9% of women discontinued supplementation, most commonly due to side effects. The drop-off rate was lower during the second KAP survey conducted (21 months into the program). Key recommendations included strengthening follow-up for management of side effects for IFA and CHW-led reminders for women. For breastfeeding, traditional practices appeared entrenched, with some clients giving water during hot weather with sufficiency of breastmilk met with disbelief (“breast milk alone not enough”). Despite the quantitative adoption data indicating positive trends, traditional practices remained challenging to overcome. Practices like giving sugar water to newborns instead of colostrum from the belief that first milk was harmful or insufficient or newborns traditionally being kept separate from mothers after birth were barriers to essential newborn care practices.

### Implementation Cost

Total program cost was 831,457 USD. The costs include items such as commodities and supplies, but also estimated proportional breakdown of time committed by the CHW and program staff on each of these interventions. That amounted to a cost per participant of 714 USD, with 1165 women enrolled. This translated to 117 USD per visit when total costs were divided by total number of visits. When excluding start-up and one-time costs, the costs lower to US$678 per participant and US$111 per visit.

The main cost driver was personnel. Staff of the NGO (35%) and Non-staff Personnel such as the CHWs (20%) together accounted for 55% of total spending, with Program Supplies & Activities adding a further 19%. This reflects a staff- and service-delivery-intensive model with minimal capital costs. Because the program is delivered almost entirely through CHWs and program staff, personnel costs appear both directly (as staff lines) and indirectly as labor is embedded in each intervention. When shared support and set-up costs are distributed across interventions, spending is spread relatively evenly across the intervention specific service package as follows: general costs non-specific to an intervention, such as danger sign recognition and promotion of facility-based antenatal care ($159\comma \; 391\semicolon \; 19&percnt\semicolon \; \rpar and Handwashing & Sanitation \lpar $156,283; 19%) are the largest, followed by Malaria ($129,871; 16%), with the remaining components each around 11–12%.

### Penetration

Supervisors and CHWs supported linkages to facility-based services through referrals and client education on care-seeking. Facility integration was still limited by poor availability and readiness of facilities and weak referral support in the general geographic area. The referral system was supported by the program with subsidized ambulance support, but the program also did not have a formalized system to track referrals for completion.

The program had a collaboration agreement with the federal and state ministries to conduct supervision and select training sessions together. Government engagement included joint supportive supervision twice a year with the state Ministry of Health. However, there was a lack of data on if the collaboration experienced any problems and how it evolved over time. The program established a federal Technical Advisory Group, co-chaired by the Federal Ministry of Health with members from the State Ministries of Health, multilateral organizations, international Non-Governmental Organizations, research institutions, and IRC to ensure buy-in and technical guidance. A program team representative also participated in the federal Reproductive Maternal Newborn Child and Adolescent Health strategy and federal Community Health Strategy revision processes to ensure program and policy alignment.

### Sustainability

Sustainability was a cross-cutting theme particularly linked to acceptability, appropriateness, and feasibility of CBMNC. The acceptability of CBMNC among participants, CHWs and providers was often conditioned on sustainability factors. For instance, health providers highlighted the need to integrate facility level care and trained professionals. CHWs also shared similar perspectives, emphasizing their motivation to continue providing care beyond this program, through integration into a strengthened health system. The systemic and structural challenges identified, such as poor facility linkage and CHW support and skills imply a health system strengthening approach is needed in future to sustain positive outcomes. While the program did establish some links to the formal health system, such as ambulance support for referrals, the data suggests that more investment is needed to sustain these linkages. And while strong relationships existed between program implementers and Ministry of Health, a concrete plan would be needed to ensure ownership by the latter. The data also highlights community expectations that extend beyond the postnatal duration of the program design; the termination of service delivery prior to the length desired by the community appeared to reduce trust and satisfaction.

## Discussion

Existing research suggests that community-based maternal and newborn health services can meaningfully reduce morbidity and mortality, yet much of this evidence stems from highly controlled research settings (20). This implementation research study contributes valuable “real world” evidence to guide similar initiatives in resource-limited and humanitarian contexts, including the future expansion of Somalia’s national *Marwo Caafimaad* program. The model showed potential to improve access to key MNH services in rural areas where facility-based care is scarce, though evidence on uptake directly attributable to the model was mixed. As seen in other settings, CHWs were widely accepted and trusted by women and communities, particularly when selected through locally participatory processes (21), and the all-female cadre, as intended in *Marwo Caafimaad,* likely contributed to the high acceptance. Participants and stakeholders perceived the model to be a reasonable epidemiological, sociocultural, and contextual fit for the setting. However, the findings highlight that planning for scale-up must address a range of operational considerations, including workload, visit frequency, household geographic dispersal, referral pathways and facility linkages, literacy-appropriate data systems, community engagement, targeted equity strategies, and long-term financing. Taken together, these findings indicate that successful scale-up will require attention not only to the content of the CBMNC package, but equally to strengthening the delivery, monitoring, and adaptive systems needed to implement it effectively and equitably.

### Burden of home visits and implications for CHW role

CHWs chose to move away from a visit schedule originally designed around stages of pregnancy, with fewer visits in early pregnancy and more frequent visits as delivery approached, adopting instead a consistent monthly schedule. This shift was driven by logistical challenges, particularly the difficulty of coordinating home visits alongside non-routine tasks such as enrolling newly pregnant women into the program and distributing the second 60-day course of IFA supplements. This suggests that feasibility was shaped not only by the overall volume of visits expected, but also by the complexity of managing multiple tasks operating on different and overlapping timelines. Notably, the decision to take on a higher number of visits was made despite CHWs’ own expressed concerns about workload, which may reflect the intrinsic motivation of selected CHWs to serve and support their communities.

Given the high mobile penetration in Somalia, technological solutions to home visit planning could be within reach. At the simplest, many mobile applications already exist around planning and scheduling (22), but with new technology emerging, there may be ways to optimize when and which home visits to make based on the location of homes using GPS, without major sacrifices to timing or frequency designed around CHW literacy, network coverage, language, and field usability, so that it reduces rather than adds to CHW workload. For instance, a study from Madagascar attempted to optimize on-foot routes for CHWs using mapping software (23). There may also be ways to set up an algorithm to alter home visits to anticipated disruptions like seasonal weather patterns or Ramadan, when both participant and provider have less capacity. These adaptations, particularly automated ones, to cadence or quantity of visit need to be conducted while complementing the strength of the CHW model; CHWs were seen as trusted individuals who respected cultural and religious norms in what and how they delivered the services, and removing or diminishing the human elements of decision-making on visit routines could potentially jeopardize what makes the CHWs effective.

Relatedly, sufficiency of CHW density also needs to be revisited; our program observed that even altering from a 1:150 (per *Marwo Caafimaad* program) to 1:50 CHW-to-household ratio appeared insufficient. Acknowledging that our observations are taken from a program was only piloted in one specific location under one implementer, a move away from a “one size fits all” rule of one CHW per a set number of households may benefit future scale-up, with an example from one study which used geospatial mapping to identify high need areas to increase geographical coverage (24). For *Marwo Caafimaad* scale-up, CHW density may need to account for household dispersion, travel time, seasonal mobility, visit frequency, commodity distribution responsibilities, and reporting burden, rather than relying only on the number of households assigned to each CHW. In addition, it is important to consider any future task-shifting for other MNH promotion activities that may add to the scope of CHW work while maintaining the same infeasible ratio. A systematic review showed that while a ratio of one to 100-200 households was common in many contexts, the CHWs who expressed satisfaction in their work at these ratios also had adequate supervision, remuneration and equipment (25).

### Community engagement

The program drew on multiple community engagement pathways to reduce barriers to uptake. These platforms appeared to play an important role in introducing the program, easing initial skepticism and building trust in CHWs. However, the available data did not permit a meaningful comparison across these pathways; little insight could be drawn from available data on which engagement platforms contributed most to positive implementation outcomes, or what the relative time and cost implications of each approach were. This makes it difficult to determine the minimum effective combination of engagement strategies needed for routine implementation. Much of the existing literature supports peer groups, including group ANC, as effective in improving behaviors or health outcomes such as breastfeeding, perinatal mental health and some on direct newborn health outcomes (26–28). But the evidence base of such groups that bring mothers or fathers together are for highly structured programs. Future models should specify what each engagement pathway is expected to change, such as trust, attitudes, and self-efficacy, and determine what community engagement pathways make the most impact on those intermediary outcomes to uptake.

### Complementarity of community and facility services

As a pilot program, there was limited investment made in the integration of the CBMNC model into the facility-based health system. Our data suggests that community awareness alone did not lead to increase in facility-based service uptake, as women continue to face persistent challenges in transportation, referral completion, facility preparedness, and the availability of skilled healthcare providers. Existing evidence on how to increase institutional delivery rate points to factors such as availing demand-side financing (e.g. conditional cash transfers, vouchers (29)) and increasing quality of facility-based services including health workforce availability (30, 31). In addition, Somalia would need to address the general scarcity of facilities; the national health infrastructure density is reported at 0.9 facilities per 10,000 population, while Galmudug, where the program was implemented, has been reported at 0.58 facilities per 10,000 population (32), both below the WHO SARA service-availability benchmark of 2 facilities per 10,000 population (33). Given records of overwork of existing midwives in this context (34), task shifting of elements like antenatal and postnatal counseling and commodity distribution to CHWs may allow for services that require a trained clinician, like support during labor and delivery, to be executed with effective coverage.

### Policy and financing barriers

In the context of Somalia, there were policy gaps between the evidence base and allowable or operationalized interventions in the country. The final pilot package was shaped not only by potential mortality impact, but also by policy permissions, commodity availability, training feasibility, and financing constraints. Side effects for IFA were one of the most salient issues raised by women throughout the program; multiple micronutrient supplements have already been demonstrated to have greater pregnancy benefits (35) and lower side effects over IFA (36), but is yet to be listed on Somalia’s Essential Medicines List to the best of our knowledge and many barriers to scaling remain (37). Misoprostol was an intervention determined by the constrained optimization model as highly life-saving in the context of Somalia but was removed from the final list of interventions due to the current policy not allowing for community-based distribution. These examples show that optimizing a community-based MNH package requires more than identifying evidence-based interventions; it also requires policy alignment, supply-chain readiness, supervision capacity, and safeguards for safe delivery at community level.

The exclusion of these evidence-based interventions in optimizing health impact is also compounded by low domestic financial investment in the health system, which worsens resource allocation for the community health system. A political economy analysis on prioritization of MNH in Somalia noted community-level service delivery as a major MNH gap with fluctuation of international donor or actor interest hampering meaningful and sustained support, financial or otherwise, for community-based services, with one respondent noting, “When the project ends, community health workers lose their jobs. Unless we have domestic reliance and do not depend on donor financing, we will not have a sustainable MNH improvement (38).” Stakeholders view increased government ownership as central to strengthening maternal and newborn health prioritization, advancing policy implementation, and tackling health system challenges in Somalia. Embedding policy delivery within the federal structure is an emerging and growing ambition for the country (39). Greater advocacy is needed to encourage international actors to align with this direction, fostering more sustainable, long-term systems strengthening for mothers and newborns in Somalia.

A recent review on costs of CHW programs on MNH reported that 57 studies reported a cost range from $0.19 to $1,547 per beneficiary (40), putting the cost from this model to be the middle of that range, with likelihood that the lower costs reported in the review have not accounted for the full operational costs of a program like our analysis did. Also, across seven CBMNC programs costed in a multi-country analysis, costs ranged widely (Bolivia’s program cost $258 per mother-baby pair versus Tanzania’s $19), which was attributed to the Bolivian CHWs operating in remote highland communities serving fewer mothers (41). Spreading of fixed costs across a small client base could also be leading to higher cost efficiency in the rural Somalia context. The cost from our analysis also does not take into account either real or opportunity cost saved in the facility-based system, with supplementation of services that either are not or cannot be provided at the health facility, as noted in our data, or reduction of facility overcrowding. While there may be promise of reducing this specific program’s per-unit costs as the model scales, some literature suggests that there are instances in which diseconomies of scale also happen due to costs incurred specifically for scale-up (42). More scenario planning based on the pilot experience should be conducted to best identify the level of scale that optimizes cost efficiency.

### Addressing low literacy

Collection and collation by CHWs of monitoring data proved to be challenging, despite adaptations made for low literacy. Further simplification may need to be considered. The challenge was not limited to data quality; complex tools increased the workload for CHWs who were already covering long distances, juggling multiple visits, distributing commodities, and following up with clients. For future scale-up, there is a need for monitoring systems that are simple enough for routine data to be used not just for reporting, but also for supervision, stock management, referral tracking, and identifying CHWs who may need extra support. Previous research has tested text-free mobile interfaces where questions would be asked of respondents via audio recording to which respondents respond by clicking color-coded boxes on a tablet (43, 44). Tools like these could be helpful, but only if they are built with CHW literacy, Somali language, network reliability, and the practical conditions of field work.

### Limitations

As a pilot implementation research project, its primary objective was to assess feasibility, acceptability, and implementation processes, rather than to generate definitive estimates of impact on maternal or newborn outcomes. Routine monitoring data faced challenges in collection and collation, even after adaptations for low literacy, potentially compromising data completeness and consistency. Although triangulation across data sources improved interpretation, certain implementation domains were less thoroughly captured, including referral completion, facility readiness, comparative performance of community engagement mechanisms, and reach among adolescents, women with disabilities, and seasonally mobile households. The research was conducted in a rural area of Galmudug, Somalia, where household dispersion, transportation limitations, and health system fragility affected implementation. Consequently, the findings may not be directly generalizable to all Somali settings, but they offer relevant insights for designing and scaling up community-based maternal and newborn care in similar low-resource and fragile contexts. There is also a risk of confirmation bias, with program implementers playing a role in the data extraction; we prioritized the value of familiarity with the data and context of the extractor over the potential for this bias to influence the results.

## Conclusion

This implementation research study examined CBMNC in resource-constrained and humanitarian settings, offering insights into implementation successes and challenges of community-based service delivery models beyond controlled research environments. The model showed promise in improving access to maternal and newborn health services in rural areas, with CHWs widely accepted by women and communities, and the intervention broadly viewed as a good fit epidemiologically, socio-culturally, and contextually. However, for successful and effective scale-up, the findings highlight the need to address practical challenges such as CHW workload, referral systems, health literacy, and equitable access; expansion must strengthen not just the content of care, but the systems needed to deliver it effectively.

## Data Availability

No data was generated by this study. Data from the primary research referred to here here will be made available on Figshare when papers from those studies are published separately.

## Acknowledgements

This research was funded by UK International Development from the UK government as part of the EQUAL Research Programme Consortium (PO 8613) and the program implementation funded by a private donor. The funders had no role in study design, data collection and analysis, decision to publish, or preparation of the manuscript. We thank Dr. Abdirisak Dalmar and Tom Kipruto Maritim for their support.

## Notes

### Competing Interest Statement

The authors have declared no competing interest.

### Author Declarations

All primary research referred to here received Institutional Review Board approval from the Somalia Ministry of Health, Somali Research and Development Institute, and the IRC respectively.

